# Loss of heterozygosity in gastric cancers in a set of Mexican patients

**DOI:** 10.1101/2024.07.29.24311063

**Authors:** Violeta Larios-Serrato, Hilda-Alicia Valdez-Salazar, Javier Torres, Margarita Camorlinga-Ponce, Patricia Piña-Sánchez, Fernando Minauro-Sanmiguel, Martha-Eugenia Ruiz-Tachiquín

## Abstract

Worldwide, gastric cancer (GC) is a common malignancy with the highest mortality rate among digestive system diseases. The present study of GC and loss of heterozygosity (LOH) is relevant to understanding tumor biology and establishing essential aspects of cancer. Here, DNA samples from Mexican patients with diffuse GC (DGC), intestinal GC (IGC), or non-atrophic gastritis (NAG; control) were purified, and whole-genome high-density arrays were performed. Posteriorly, LOH was identified among the tissue samples, and cancer genes and signaling pathways were analyzed to determine the most altered. Detailed bioinformatics analysis was developed to associate LOH with the Hallmarks of Cancer according to their frequency in patient samples, participation in metabolic pathways, network interactions, and enrichment of Cancer Hallmark genes. LOH-genes in GC were PTPR, NDUFS3, PAK3, IRAK1, IKBKG, TKTL1, PRPS1, GNAI2, RHOA, MAPKA, and MST1R. Genes that stand out at NAG involve proliferation and growth; those at IGC trigger genomic instability, tissue invasion, metastasis, and arrest of cell death; and those at DGC involve energy metabolism, the destruction of immune evasion, and replicative immortality. Other events, such as sustained angiogenesis, were similar between NAG-IGC-DGC. Together, these are molecular, cellular, and metabolic events that must be monitored in GC patients. Our findings must be validated to develop molecular tests for diagnosis, prognosis, treatment response, and, most importantly, screening tests.

## Introduction

Gastric cancer (GC) ranks fifth in the world according to incidence and mortality (1). GC is a multifactorial disease with environmental and genetic factors impacting its occurrence and development. The GC incidence rate rises progressively with age; the average age for diagnosis is 70 years, a small portion of gastric carcinomas (10%) are detected at younger ages (45 or less) and becomes good chances to look for GC early genetic alterations or carcinogenesis pathways since those patients are less exposed to environmental facts. Carcinogenesis is a multistage process with a progressive development that involves gene mutations and epigenetic alterations (2).

GC molecular classification involves four subtypes: tumors with chromosomal instability (CIN), tumors with microsatellite instability (MSI), genomically stable tumors (GS), and EBV-positive tumors (3). CIN is one of the significant genomic instability pathways involved in gastric carcinogenesis, and it is characterized by losses or gains of whole chromosomes that result in aneuploidy. CIN may also involve changes in portions of chromosomes, which include allelic losses like loss of heterozygosity (LOH), gene deletions, amplifications (4, 5) or rearrangement (6).

Also, two main GC histotypes are recognized: intestinal and diffuse. Although most of the described genetic alterations have been observed in both types, different genetic pathways have been hypothesized. Genetic and epigenetic events, including 1q LOH, microsatellite instability, and hypermethylation, have mostly been reported in intestinal-type gastric carcinoma (IGC) and its precursor lesions, whereas 17p LOH mutation or loss of E-cadherin are more often implicated in the development of diffuse-type gastric cancer (DGC) (7).

LOH has been identified as an etiological factor in chromosomal instability, which promotes the progression of various types of cancer, including GC (6). LOH involves the loss of one of the two gene alleles in a cell, which can lead to the inactivation of tumor suppressor genes and contribute to the development and progression of cancer. There are two types of LOH: (1) copy number loss LOH (CNL-LOH), which implies the loss of alleles, for tumor suppressor gene as an example, and (2) copy number neutral LOH (CNN-LOH), without any affecting function nor contributing to the disease development (8). A complete or partial deletion of a chromosome leads to CNL-LOH, while CNN-LOH is mainly caused by acquired uniparental disomy (UPD) and genetic conversion and occurs without a net change in copy number (9) (10).

Various molecular techniques have been used to investigate the role of LOH in GC (11), such as polymerase chain reaction (PCR), microsatellite marker sites PCR (12), multiplex ligation-dependent probe amplification (MLPA) (13), polyacrylamide gel electrophoresis (PAGE) (14), silver stain (15), exome sequencing (16), Illumina (17) and Affymetrix (18) microarrays. The identification of LOH-events can be assessed by gene expression using RNAseq and RT-PCR (19) and protein expression with immunohistochemistry (IHC) (18). LOH has also been correlated with CpG hypermethylation processes in patients with GC (20). The LOH-genes associated with CG most frequently reported are TP53 (21), PTEN (16), RB1, and BRCA1 (22). The loci involved in tumor suppression are located on chromosomes 1, 3, 7, 8, 11, 12, 13, 18, and 22 (17, 23). Different scores have also been proposed to establish risk or diagnosis based on LOH (22). For the above, our interest arose in determining the LOH patterns in a group of IGC, DGC, and non-atrophic gastritis (NAG) samples to find guidelines for therapeutic targets and data that enrich the knowledge of cancer biology.

## Material and Methods

### Samples

Tissue samples were obtained from 21 patients (5 females and 16 males) that met the criteria for diffuse gastric cancer (DGC, n = 7) and intestinal gastric cancer (IGC, n = 7) diagnoses, and subjects with non-atrophic gastritis (NAG, n = 7) as controls. To guide the investigation of relevant alterations, the present analysis focused on LOH-events present in at least three patients (cut-off, ≥ 3 patients; ≥ 40% samples) to identify the most relevant GC alterations (Table 1). The DNA extraction, DNA quality assessment, and high-density whole-genome processing microarray analysis were done according to Larios *et al.* 2022 (24).

**Table I.** Research questions and their rationale.

| ID | Sex | CT | %CC | *H. pylori* | TNM | Treatment |
| --- | --- | --- | --- | --- | --- | --- |
| 1 | M | Intestinal | 70 | Positive | I B T1 N1 M0 | Naïve |
| 2 | M | Intestinal | 60 | Negative | IIA T4 N0 M0 | Naïve |
| 3 | M | Intestinal | 70 | Negative | IIA T3 N2 M0 | Naïve |
| 4 | F | Intestinal | 60 | Negative | IV T4 N2 M0 | Naïve |
| 5 | M | Intestinal | 50 | Negative | II T3 N0 M0 | Naïve |
| 6 | M | Intestinal | 60 | Negative | IIB T3 N2 M0 | Naïve |
| 7 | M | Intestinal | 60 | Negative | III A T4 N0 M0 | Naïve |
| 8 | M | Diffuse | 70 | Negative | IV T4 N3 M0 | Naïve |
| 9 | M | Diffuse | 70 | Negative | III A T2 N3 M0 | Naïve |
| 10 | M | Diffuse | 70 | Negative | II T1 N0 M0 | Naïve |
| 11 | M | Diffuse | 60 | Negative | IV T4N2M1 | Naïve |
| 12 | M | Diffuse | 60 | Negative | IV T4N2M0 | Naïve |
| 13 | M | Diffuse | 50 | Negative | IV T4, N2 M0 | Naïve |
| 14 | M | Diffuse | 50 | Negative | IV T4 N2 M0 | Naïve |
| 15 | M | NAG | 0 | Negative | NA | NA |
| 16 | M | NAG | 0 | Negative | NA | NA |
| 17 | F | NAG | 0 | Negative | NA | NA |
| 18 | M | NAG | 0 | Positive | NA | NA |
| 19 | F | NAG | 0 | Positive | NA | NA |
| 20 | F | NAG | 0 | Positive | NA | NA |
| 21 | F | NAG | 0 | Negative | NA | NA |

ID, identification code; CT, cancer type; CC, cancer cells; M, male; F, female; NA, not applicable; TNM, T: the extent of the primary tumor; N: the absence or presence and extent of regional lymph node; M: the absence or presence of distant metastasis (24). NAG, non-atrophic gastritis.

### LOH processing

The raw intensity files (.CEL) retrieved from the commercial platform Affymetrix® CytoScan™ microarray (Affymetrix; Thermo Fisher Scientific, Inc.), were analyzed using Chromosome Analysis Suite (ChAS) v4.3.0.71. The construction of the GRCh38 genome (December 2013) was used as a reference model and CytoScanHD_Array.na36.annot.db file for annotation. Data processing was based on the segmentation algorithm, where the Log2 ratio for each marker was calculated relative to the reference signal profile. To calculate the LOH, the data were normalized to baseline reference intensities using ChAS reference model including 284 HapMap samples and 96 healthy individuals. The Hidden Markov Model (HMM) was used to determine the LOH segment calls. The customized conditions were filtered to determine LOH, 3 Mb, and 50 Single Nucleotide Polymorphisms (SNPs). The Median Absolute Pairwise Difference (MAPD) and the Single Nucleotide Polymorphism Quality Control (SNPQC) score were used as the quality control parameters. Only samples with values of MAPD < 0.25 and SNPQC > 15 were included in the further analysis.

### Bioinformatics analysis

To generate a list of genes and frequencies for altered regions, Practical Extraction and Report Language (Perl) scripts (25) were developed to load the LOH segment data files generated by ChAS 4.3.0.71 for each sample, including chromosomes and cytogenetic bands and Online Mendelian Inheritance in Man information, and haploinsufficiency predictions version 3 information from the DatabasE of Genomic Variation and Phenotype in Humans, using Ensemble Resources (DECIPHER v11.25). Custom scripts were developed in Perl v5.32 to obtain the frequency of LOH -genes and -cytobands and the length of events.

The genes altered in at least three patients (cut-off, ≥ 3) with DGC, IGC, and NAG were included for analysis and visualization. The genes were compared by generating Venn diagrams with the Jvenn server (26). Cancer Hallmarks enrichment analysis (*p*.adjust < 0.05) was performed with a collection of 6,763 genes (https://cancerhallmarks.com/). The results were reviewed using the Catalog of Somatic Mutations in Cancer database (COSMIC v100) (27) the Hallmarks of Cancer database (HOCdb database) (28).

Reactome v88 performed a metabolic pathway enrichment analysis (29), considering those results significant with values less than 0.05 in the false discovery rate (FDR).

Finally, an interaction network was generated based on metabolic pathways and genetics, as well as physical and functional associations to establish the Cancer Hallmarks associated with the profile of LOH-genes IGC, NAG, and “core” IGC-DGC-NAG. Furthermore, these were determined using the STRING v12.0 prediction server (30) and Cytoscape v.3.10.0 (31), including manual annotation of their corresponding the Cancer of Hallmarks (adhesion, angiogenesis, inflammation, migration, metastasis, morphogenesis, proliferation, and survival).

## Results

### Sample characteristics

This study included tissue samples from 21 Mexican patients without treatment (naïve). Patient samples included seven DGC cases, seven for IGC, and seven more corresponding to NAG (as controls). The .CEL files and their raw intensity values obtained from the microarrays were deposited in the Center for Biotechnology Information (NCBI), with the accession key GSE117093 and BioProjet PRJNA481039.

Table 1 shows the general characteristics of the 21 patient samples, age (mean ± SD, 59.61 ± 15.94 years), sex (Female 23.8% and Male 76.2%), and the percentage of neoplastic cells for tumor tissues ranging between 50 and 70%. One IGC patient and three with NAG were positive for *H. pylori*. Data from the Tumor size, Number of nodes, and Metastasis (TNM) classification system are presented.

### Genomic detection of LOH

The LOH of the patients was estimated using the analysis described before, a meticulous process based on regions where the preponderance of SNPs does not display heterozygosity. Table 2 shows a summary of the chromosomes with the highest involvement frequency concerning the number of events (coincidences) at the LOH-regions but not strictly perfect in the chromosomal coordinates.

**Table II.**
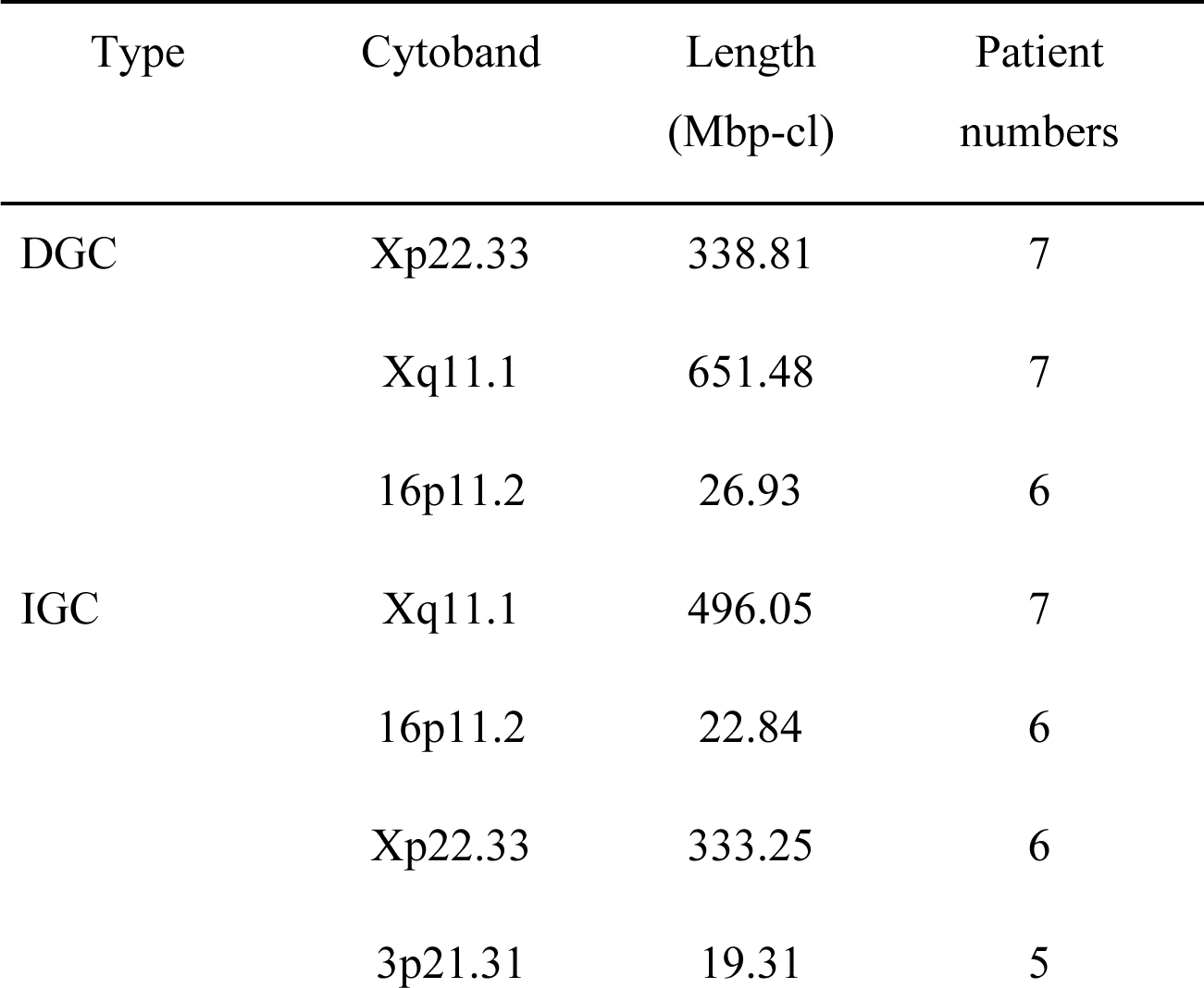
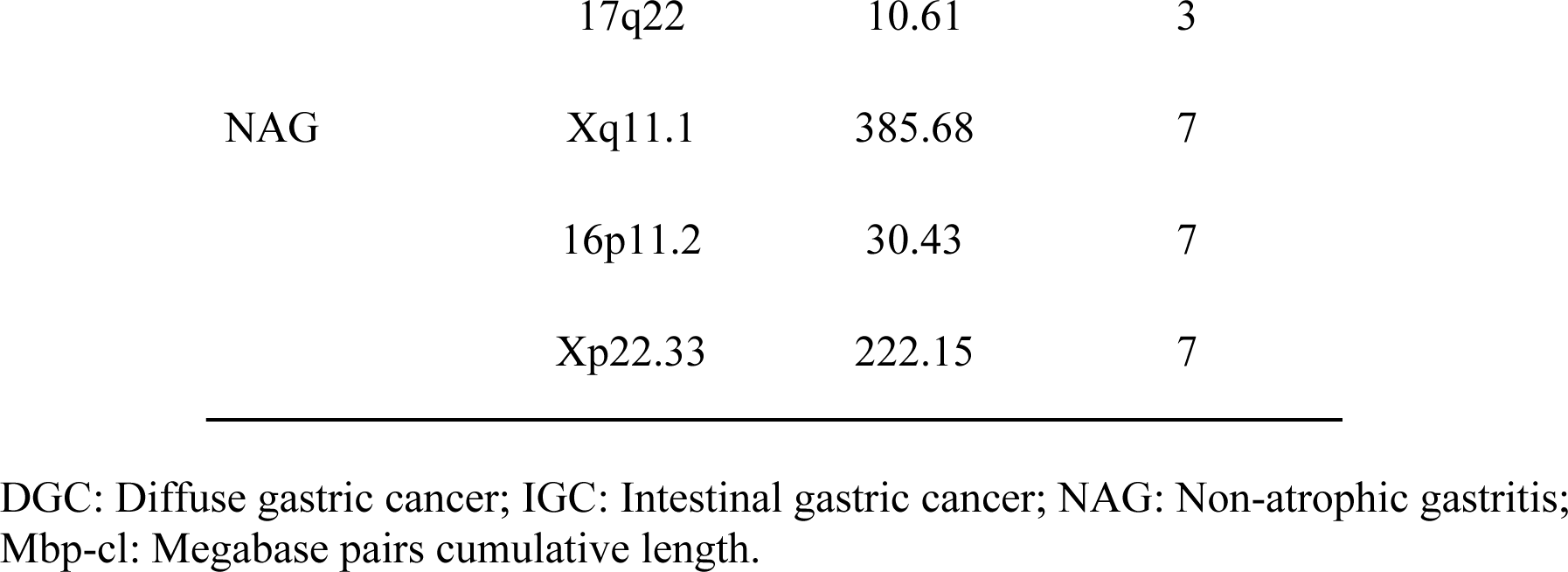
Top of altered cytobands in diffuse gastric cancer, intestinal gastric cancer, and non-atrophic gastritis.

| Type | Cytoband | Length<br>(Mbp-cl) | Patient<br>numbers |
| --- | --- | --- | --- |
| DGC | Xp22.33 | 338.81 | 7 |
|  | Xq11.1 | 651.48 | 7 |
|  | 16p11.2 | 26.93 | 6 |
| IGC | Xq11.1 | 496.05 | 7 |
|  | 16p11.2 | 22.84 | 6 |
|  | Xp22.33 | 333.25 | 6 |
|  | 3p21.31 | 19.31 | 5 |
|  | 17q22 | 10.61 | 3 |
| NAG | Xq11.1 | 385.68 | 7 |
|  | 16p11.2 | 30.43 | 7 |
|  | Xp22.33 | 222.15 | 7 |

DGC: Diffuse gastric cancer; IGC: Intestinal gastric cancer; NAG: Non-atrophic gastritis; Mbp-cl: Megabase pairs cumulative length.

Our data, which includes the megabase pairs cumulative length (Mbp-cl) of our tissue samples, were also reviewed (Table S1), the LOH-gene frequency data, chromosomes, and cytobands, is presented. Table S2 displays the accumulated LOH-length (Mb) values per chromosome, to determine if more extended losses indicate more damage.

In DGC patients, the affected chromosomes with Mbp-cl and the specified number of LOH-events were 6, 8, 16, and X; at IGC, they were chromosomes 3, also 16, and 17. Chromosomes 6 and 8 are associated with DGC, while 3 and 17 are associated with IGC (Table 2 and S2). Following, we found that there are 3,361 LOH-genes in DGC (Table S1); chromosomes Xq11.1/Xp22.23 in 7/7 male patients and chromosome 16p11.2 in 6/7 male patients (Tables 3 and S3) were the most altered.

In IGC, 2,490 LOH-genes were determined (Table S1) with chromosomes Xq11.1 for 7/7 patients, Xp22.33 for 6/7 patients, 16p11.2 for 6/7 patients, 3p21.31 for 5/7 patients and 17q22 for 3/7 patients (Table 3 and Table S3) as the most altered.

**Table III.** Top of altered cytobands in diffuse gastric cancer, intestinal gastric cancer, and non-atrophic gastritis.

| Type | Cytoband | Length (Mbp-cl) | Patient numbers |
| --- | --- | --- | --- |
| DGC | Xp22.33 | 338.81 | 7 |
|  | Xq11.1 | 651.48 | 7 |
|  | 16p11.2 | 26.93 | 6 |
| IGC | Xq11.1 | 496.05 | 7 |
|  | 16p11.2 | 22.84 | 6 |
|  | Xp22.33 | 333.25 | 6 |
|  | 3p21.31 | 19.31 | 5 |
|  | 17q22 | 10.61 | 3 |
| NAG | Xq11.1 | 385.68 | 7 |
|  | 16p11.2 | 30.43 | 7 |
|  | Xp22.33 | 222.15 | 4 |

DGC: Diffuse gastric cancer; IGC: Intestinal gastric cancer; NAG: Non-atrophic gastritis; Mbp-cl: Megabase pairs cumulative length.

Finally, in NAG 4,748 LOH-genes were determined (Table S1). Chromosomes Xq11.1 for 7/7 patients, 16p11.2 for 7/7 patients, and Xp22.33 for 4/7 patients (Table 3 and Table S3) were the most altered.

Interestingly, LOH lengths do not look relevant for carcinogenesis, and 5-10 Kbp LOH-lengths were more common and frequent in DGC, IGC, and NAG (Table S4).

To identify the most relevant LOH in GC and NAG, we analyzed alterations occurring in at least three patients (cut-off ≥ 3). We found a similar pattern for total LOH, with more events in DGC (1157), IGC (1361), and NAG (1184). In addition, DGC had the highest number of genes affected in all samples, 7/7 (1132), followed by IGC (261) and NAG (34) (Table S1).

### Gastric cancer genes associated with LOH

A Venn diagram was constructed to examine the LOH-GC-relevant genes of at least three patients (cut-off ≥ 3) of the DGC, IGC, and NAG. We determined 1153 shared LOH-genes between DGC-IGC-NAG. IGC had 207 unique affected genes, while NAG only showed 28 (Figure 1A and Table S5). From each subset (Figure 1A), those genes with matches according to the Cancer Hallmarks Genes database, a comprehensive resource that includes 6,763 genes, are shown 241 LOH-genes were found in DGC-IGC-NAG; IGC had 55 affected unique genes and 13 genes in NAG.

**Figure 1.**
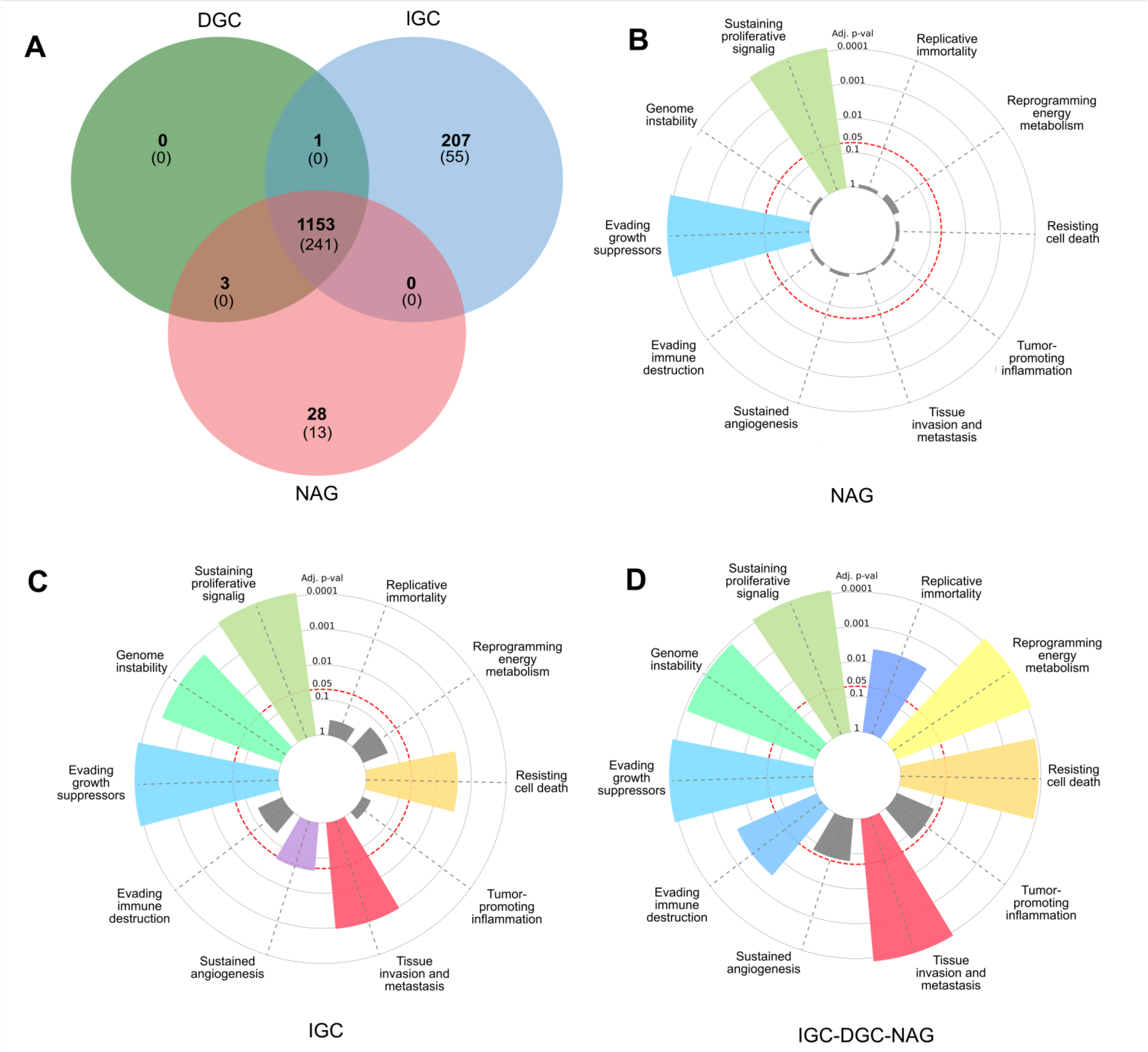
Profile of LOH-genes in gastric cancer from ≥ 3 patients. (A) The Venn diagram presents frequencies of specific and shared genes in diffuse gastric cancer (DGC), intestinal gastric cancer (IGC), or non-atrophic gastritis (NAG) in LOH. Each subset of the Venn diagram in parentheses indicates the Cancer Hallmark genes. (B, C, and D) In the radial graphs, each bar represents a Hallmark and the height of the *p*.adjust value of the category enrichment; the grays have no significance. The red dotted line represents the significance cut-off value (*p*.adjust < 0.05).

Figures 1B-D represent the enrichment of the Hallmarks of Cancer. The 241 common genes DGC-IGC-NAG present a greater number of Hallmarks than NAG, which showed fewer.

### Functional pathway analysis

Using the LOH-genes-Hallmarks identified in each subset, metabolic pathways were predicted in Reactome using *Homo sapiens* as a model organism; in Table 4, those significant metabolic pathways (*p*-value < 0.05) and a brief general description are reported.

**Table IV.**
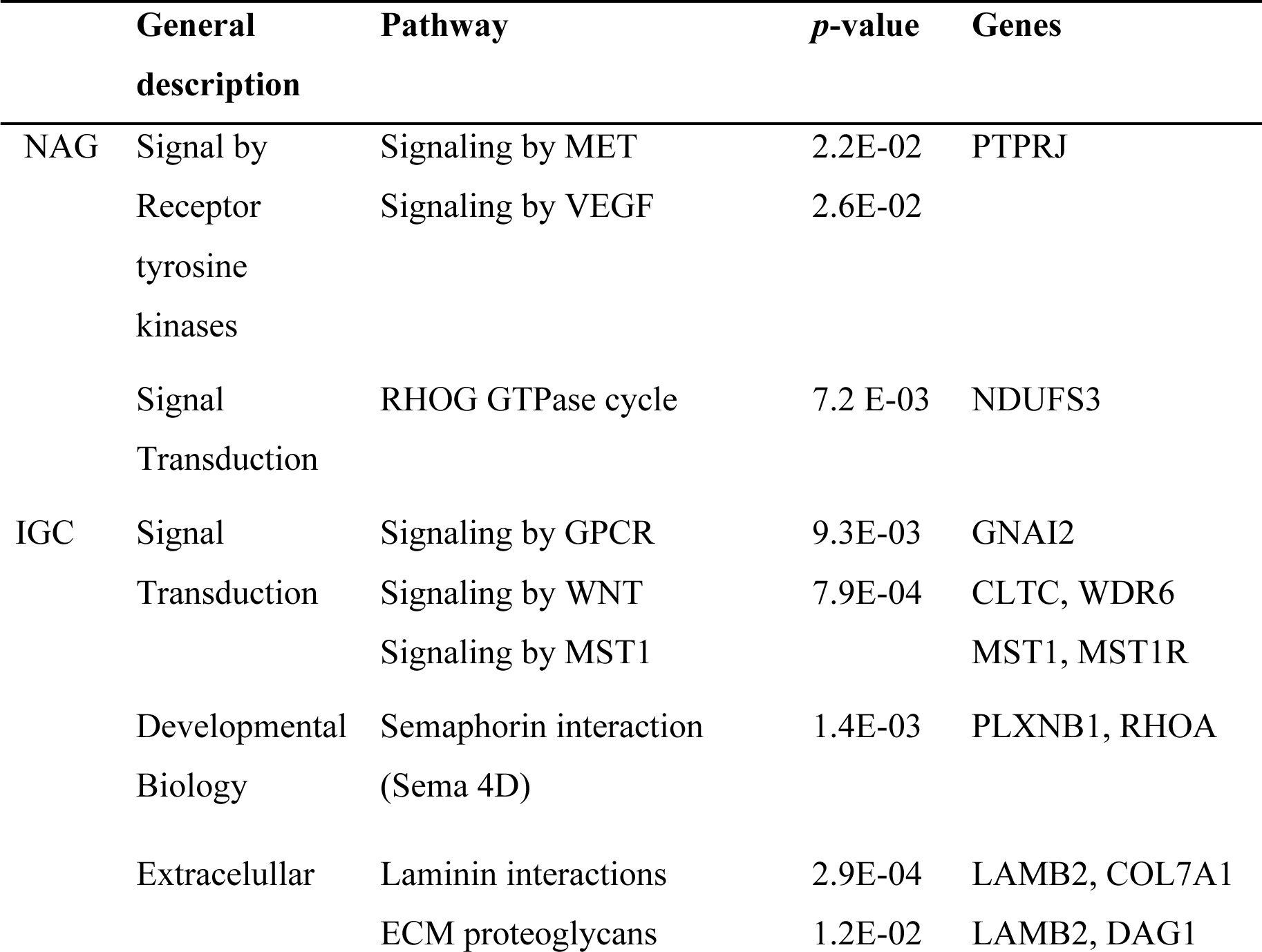
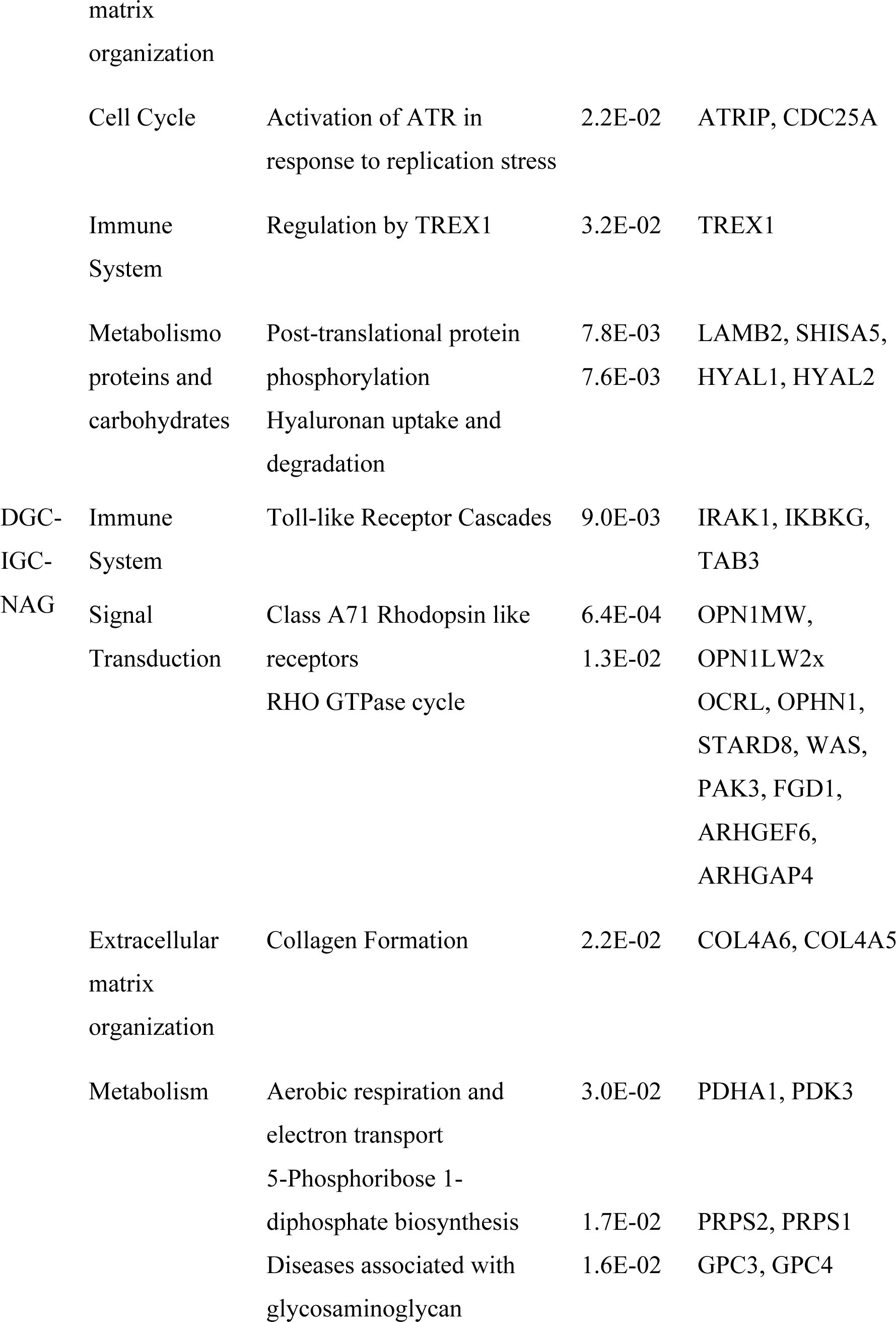
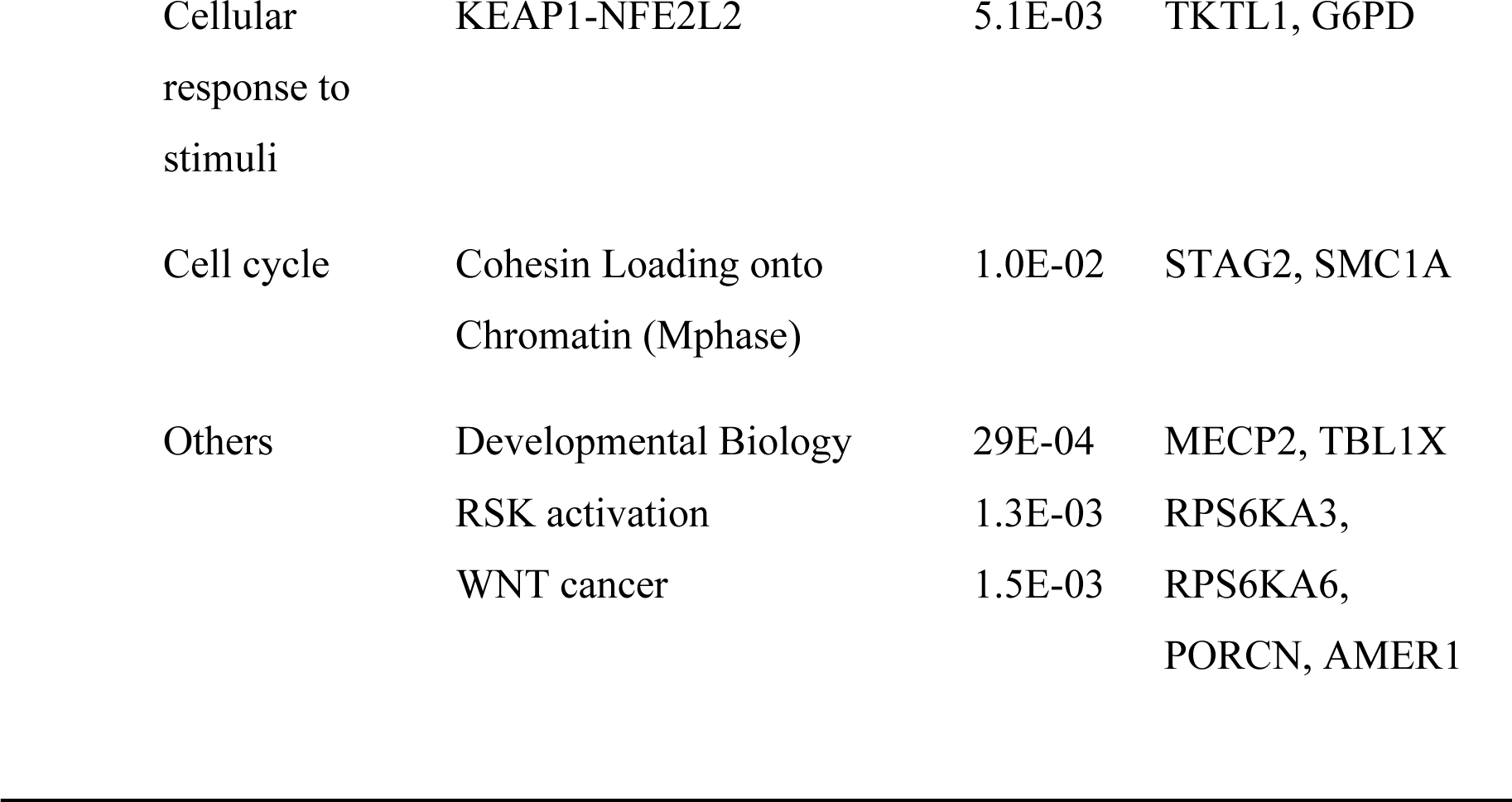
Metabolic pathways and genes-hallmarks of cancer.

### Correlation genes network

Cancer LOH-genes-Hallmarks associated with metabolic pathways were used to construct interaction networks (String, Figure 2). The connecting lines indicate associations by metabolic pathways, expression, localization, inferred interaction, genetic interactions, data mining, and neighborhood. Likewise, each node can be related to flags (events) related to report the Hallmarks of Cancer.

**Figure 2.**
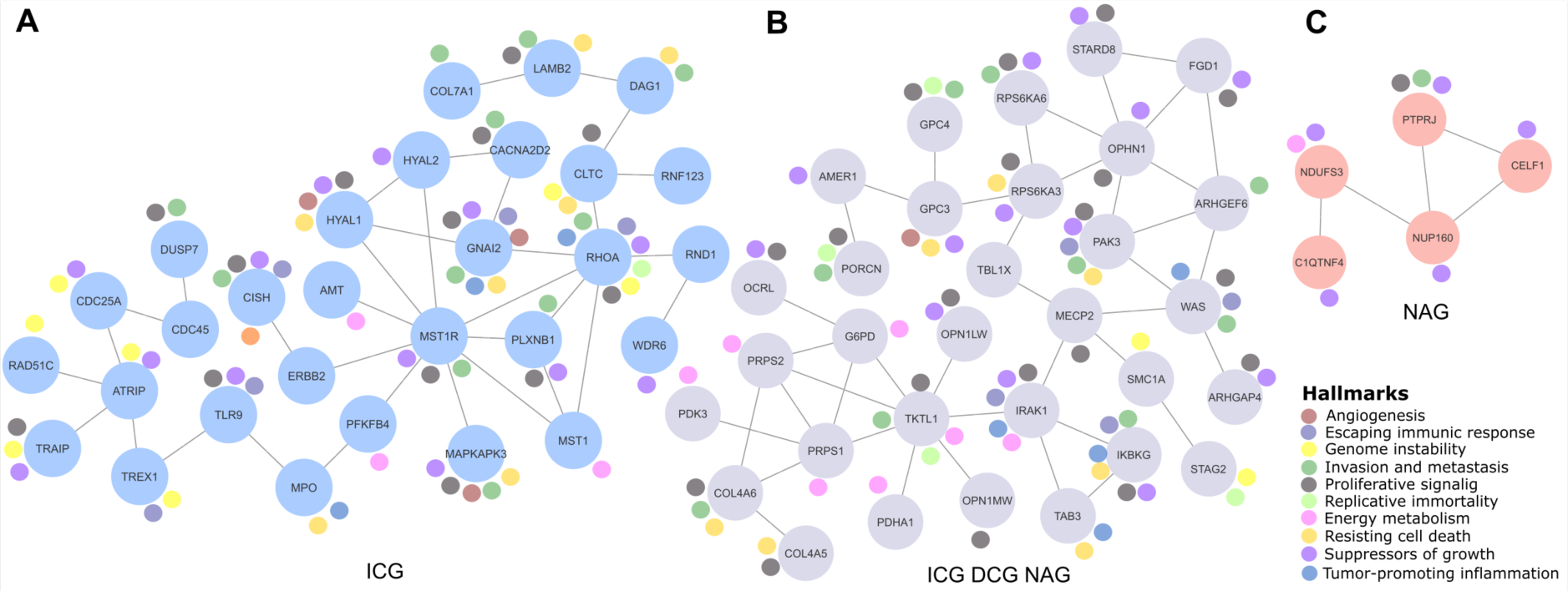
Relevant gene networks. The Hallmarks of Cancer and metabolic pathways. Ten Hallmarks are shown in the legend. The IGC-DGC-NAG network is indicated in purple, NAG in pink, and IGC in blue.

In this way, the network formed among IGC-DGC-NAG had 31 nodes; the genes with the highest number of Hallmarks were PAK3, IRAK1, and IKBKG, and the genes with the highest number of connections were OPHN1, WAS, TKTL and PRPS1. In the ICG network, there were 29 nodes. The genes associated with Cancer Hallmarks were GNAI2, RHOA, MAPKAPK3, HYAL1, and CISH, while the most connected were RHOA, MST1R, and ATRIP. Finally, the network corresponding to NAG had five nodes, PTPRJ had the most significant number of Hallmarks, and the most connected was NUP160 (Table 5).

**Table V.**
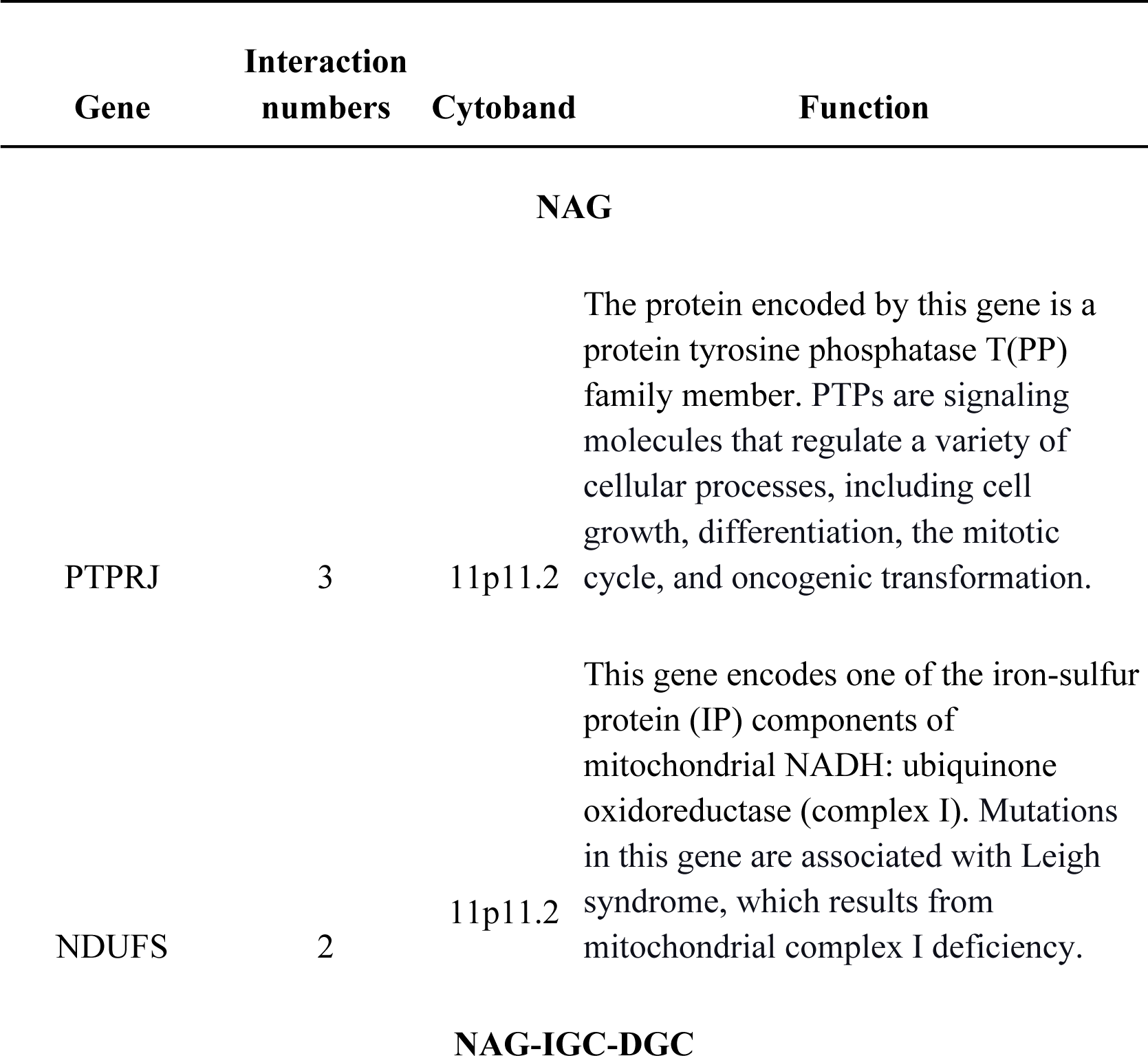
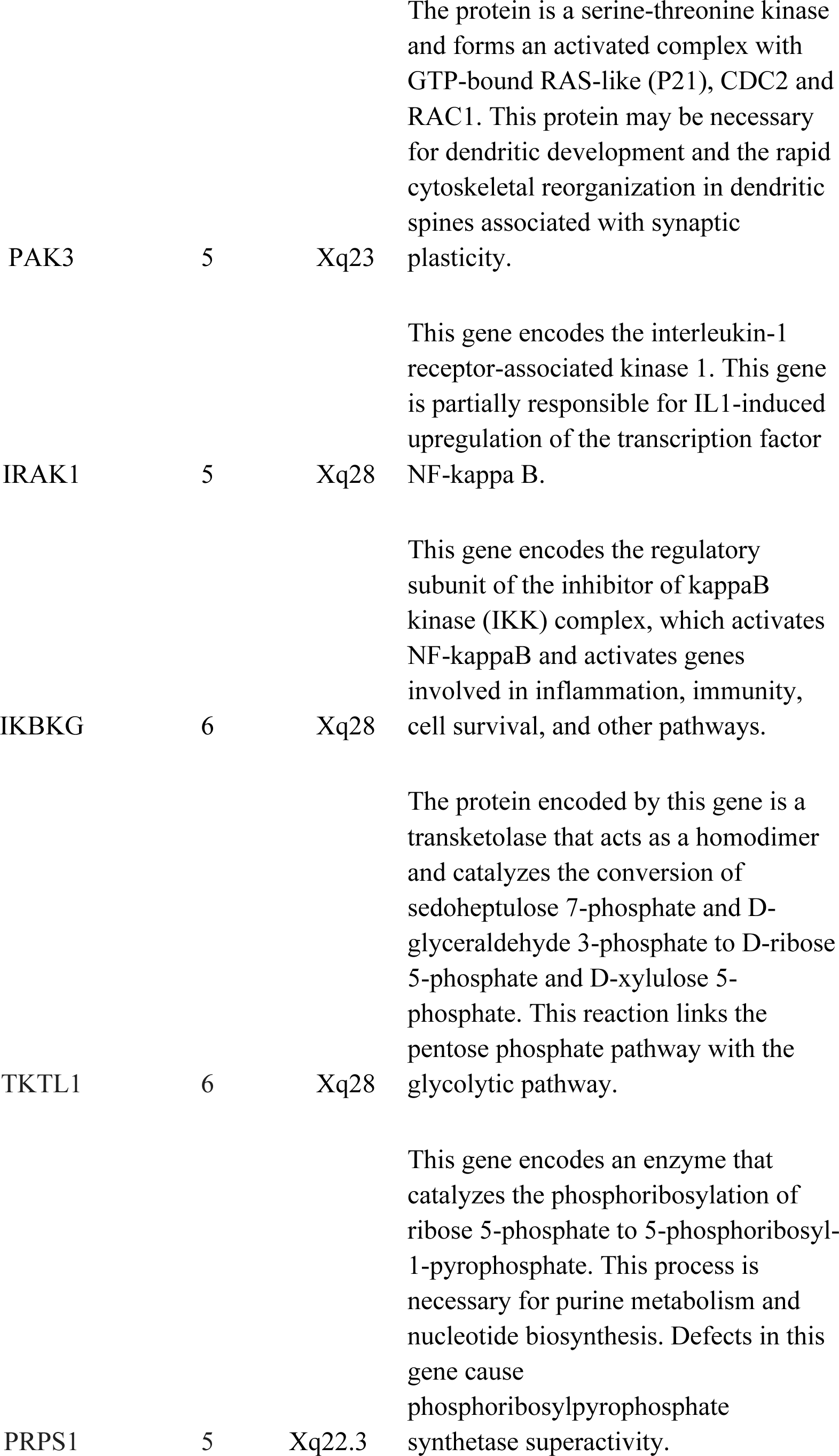
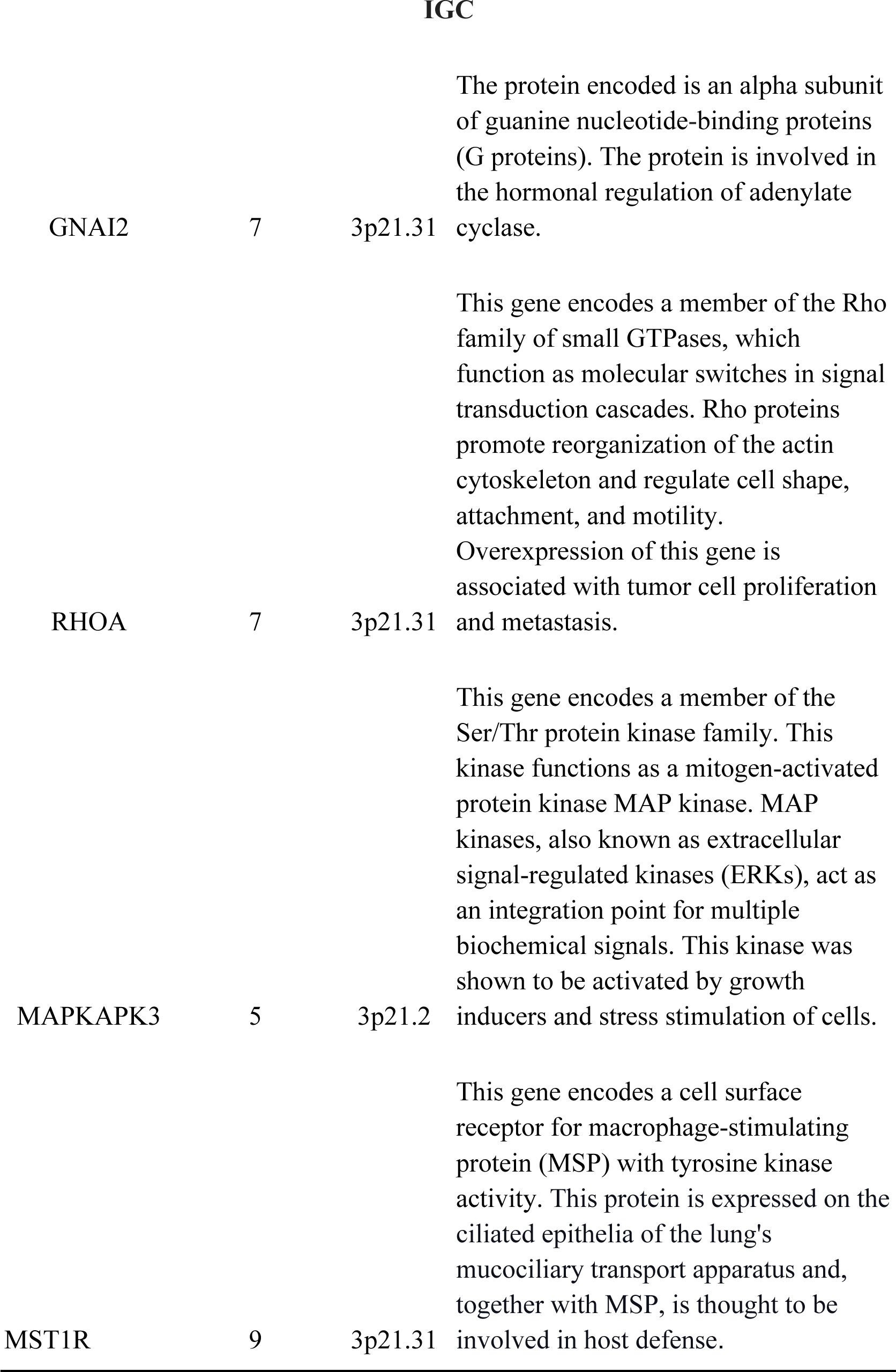
Top genes linked to the highest number of the Hallmarks of Cancer and genes detected in the stomach.

| Gene | Interaction numbers | Cytoband | Function |
| --- | --- | --- | --- |
| **NAG** |  |  |  |
| PTPRJ | 3 | 11p11.2 | The protein encoded by this gene is a protein tyrosine phosphatase T(PP) family member. PTPs are signaling molecules that regulate a variety of cellular processes, including cell growth, differentiation, the mitotic cycle, and oncogenic transformation. |
| NDUFS | 2 | 11p11.2 | This gene encodes one of the iron-sulfur protein (IP) components of mitochondrial NADH: ubiquinone oxidoreductase (complex I). Mutations in this gene are associated with Leigh syndrome, which results from mitochondrial complex I deficiency. |
| **NAG-IGC-DGC** |  |  |  |
| PAK3 | 5 | Xq23 | The protein is a serine-threonine kinase and forms an activated complex with GTP-bound RAS-like (P21), CDC2 and RAC1. This protein may be necessary for dendritic development and the rapid cytoskeletal reorganization in dendritic spines associated with synaptic plasticity. |
| IRAK1 | 5 | Xq28 | This gene encodes the interleukin-1 receptor-associated kinase 1. This gene is partially responsible for IL1-induced upregulation of the transcription factor NF-kappa B. |
| IKBKG | 6 | Xq28 | This gene encodes the regulatory subunit of the inhibitor of kappaB kinase (IKK) complex, which activates NF-kappaB and activates genes involved in inflammation, immunity, cell survival, and other pathways. |
| TKTL1 | 6 | Xq28 | The protein encoded by this gene is a transketolase that acts as a homodimer and catalyzes the conversion of sedoheptulose 7-phosphate and D-glyceraldehyde 3-phosphate to D-ribose 5-phosphate and D-xylulose 5-phosphate. This reaction links the pentose phosphate pathway with the glycolytic pathway. |
| PRPS1 | 5 | Xq22.3 | This gene encodes an enzyme that catalyzes the phosphoribosylation of ribose 5-phosphate to 5-phosphoribosyl-1-pyrophosphate. This process is necessary for purine metabolism and nucleotide biosynthesis. Defects in this gene cause phosphoribosylpyrophosphate synthetase superactivity. |
| IGC |  |  |  |
| GNAI2 | 7 | 3p21.31 | The protein encoded is an alpha subunit of guanine nucleotide-binding proteins (G proteins). The protein is involved in the hormonal regulation of adenylate cyclase. |
| RHOA | 7 | 3p21.31 | This gene encodes a member of the Rho family of small GTPases, which function as molecular switches in signal transduction cascades. Rho proteins promote reorganization of the actin cytoskeleton and regulate cell shape, attachment, and motility. Overexpression of this gene is associated with tumor cell proliferation and metastasis. |
| MAPKAPK3 | 5 | 3p21.2 | This gene encodes a member of the Ser/Thr protein kinase family. This kinase functions as a mitogen-activated protein kinase MAP kinase. MAP kinases, also known as extracellular signal-regulated kinases (ERKs), act as an integration point for multiple biochemical signals. This kinase was shown to be activated by growth inducers and stress stimulation of cells. |
| MST1R | 9 | 3p21.31 | This gene encodes a cell surface receptor for macrophage-stimulating protein (MSP) with tyrosine kinase activity. This protein is expressed on the ciliated epithelia of the lung's mucociliary transport apparatus and, together with MSP, is thought to be involved in host defense. |

## Discussion

### LOH, chromosomes and cytogenetics

GC has a high mortality rate due to its characteristics, such as late detection and silent progression; then, research into tumor biology is required to find early markers to carry out an opportune intervention.

LOH is among the aspects that could contribute to a timely diagnosis according to prognosis (11, 21). It has been proposed that LOH could function as independent prognostic markers (32), and those could even function as alternative targets for treatment (33).

LOH is involved in different cancer types, showing its importance as a “predisposing” factor. More than the length of affection, specific LOH-genes look more relevant, according to other studies. LOH-genes-NAG, TP53, PTEN, RUNX3 (34–36) could have a carcinogenic potential as signaling early events, of these TP53 (34) and PTEN (Li *et al*. 2005) have polymorphisms or copy number variations due to LOH-events (21, 37, 38).

Here, 11 LOH-genes were determined, which were selected according to their apparition frequency in the analyzed samples, their participation in metabolic pathways (*p*-value < 0.05), their established interactions (networks), and their enrichment in Cancer Hallmarks genes database (*p*.adjust < 0.05). Thus, PTPRJ and NUP160 were determined into NAG samples, RHOA, GNAI2, and MAPKAPK3 for ICG, and no unique or relevant genes were identified for DCG. NAG LOH-genes that are relevant to carcinogenesis participate in proliferation and growth, while those for IGC are on genomic instability, tissue invasion, metastasis, and the arrest of cell death; and DGC genes are for energy metabolism, destruction of immune evasion, and replicative immortality. Other genes were shared between IGC and NAG-IGC-DGC, whose *p* values are close and could be considered similar LOH-events since they are involved in sustained angiogenesis. On the other hand, IGC genes also promote inflammation, and although the *p* values are not significant, there was a difference in the NAG-IGC-DGC group. Then, those molecular, cellular, and metabolic LOH-alterations should be monitored in GC patients. These findings must be validated to develop tests with molecular profiles for diagnosis, prognosis, and response to treatment, as well as, most importantly, screening tests.

When analyzing the metabolic pathways associated with LOH-genes, the common ones were signal transduction, immune system, cell cycle, and extracellular matrix organization, which would be involved in early GC stages. Developmental biology is added to IGC samples because the semaphorins interactions (Table 4, genes PLXNB1 and RHOA), which have been found to function as tumor suppressors and inhibit tumor progression by various mechanisms, are found; however, also can function as inducers and promoters of tumor progression (39). Here (IGC) protein and carbohydrate metabolism genes are found, then processes such as post-translational modifications (phosphorylations) and hyaluronan metabolism (degradation proteins Hyal 1-3) can take place. Hyaluron plays a fundamental role in tissue architecture and the regulation of cellular function, which could be related to proliferation and migration since hyaluronan accumulation at the extracellular matrix and its derived fragments due to the altered expression of hyaluronidases enhance cancer progression, remodeling the tumor microenvironment (40), just like in GC (41).

### Marker genes

Based on the results above described, five shared genes, were determined among the three samples groups analyzed (IGC-DGC-NAG): IRAK1, IKBKG, PAK3, TKTL1, and PRPS1. These genes are associated with various functions of cancer development and progression. IRAK1 regulates inflammation genes at immune cells and could affect the disease development due to its possible oncogenic and immunological functions (42). IKBKG is linked to the NF-κB pathway, which is crucial for cancer since its involvement in cell survival and proliferation (43, 44). PAK3 can regulate cell growth and migration, potentially contributing to metastasis; it can also regulate Circ 0000190, a circular RNA that inhibits CG through the caspase-3, p27, and cyclin D axis (45). On the other hand, TKTL1 is associated with altered glucose metabolism in GC cells (46), moreover, it regulates other events such as proliferation, metastasis, epithelial-mesenchymal transition, resistance to chemoradiotherapy, and survival (47). Finally, the relapse-specific mutations in phosphoribosyl pyrophosphate synthetase 1 gene (PRPS1), a rate-limiting purine biosynthesis enzyme that confers significant drug resistances to combination chemotherapy in acute lymphoblastic leukemia (48), since these genes are shared, they could indicate a core of early genes given that they are also found in NAG, a non-cancerous lesion.

The NDUFS3 gene, which encodes a subunit of complex I of OXPHOS, was also identified among the shared genes. This gene plays a significant role in metabolic reprogramming, an early event in carcinogenesis that occurs in the first phases of disease development. Metabolic reprogramming is a crucial aspect of the Warburg effect, a common feature in cancer. Also, the *H. pylori* infection, like some of the NAG samples in this study, shows increased complex I, further highlighting the role of NDUFS3 (49). Furthermore, in the MKN28 and MKN45 cell lines derived from moderately differentiated tubular adenocarcinoma and undifferentiated adenocarcinoma of medullary type, also is observed a decrease in this complex I of the mitochondrial respiratory chain (50).

According to the NAG interaction network, PTPRJ and NUP160 genes could regulate cellular and molecular processes contributing to inflammation, immune response, energy and metabolism, and proliferation. PTPRJ encodes a protein from the tyrosine phosphatase family, dephosphorylates CTNND1, that is related to cell proliferation, adhesion, and migration (51, 52), also with inflammation and regulation of the immune response. NUP160 encodes for a nuclear porin complex protein that regulates the transport of macromolecules between the nucleus and the cytoplasm (53); its deregulation can modify gene expression and cell signaling. Four cancer signaling and progression genes were identified in the IGC network (Fig. 2); RHOA encodes for a protein of the Rho-GTPase family and is associated with signaling (54), regulates several cellular processes, including the cytoskeletal structure and cell adhesion (55). Therefore, it is linked to cancer progression. GNAI2 encodes the alpha subunit of the Guanine nucleotide-binding proteins (G proteins), and in stomach adenocarcinoma, it is considered a prognostic marker (56). Also, it is essential for the transduction of extracellular signals and is related to cellular processes such as growth, migration, regulation of proliferation, differentiation, and the response to external stimuli. MAPKAPK3 has been proposed as a therapeutic target (57) and participates in regulating cell stress response, proliferation, and survival; it has also been reported as an element of diagnosis, prognosis, and prediction (58). Moreover, the MST1R (or RON) gene encodes macrophage-stimulating receptor-1 or Ron family tyrosine kinase receptors, which can act in the HIF-1α and β-catenin pathways. This receptor is involved in oncogenesis by regulating cell migration, adhesion, and survival; it plays an essential role in the inflammatory response and metastasis. In this way, RHOA and GNAI2 are related to the signaling and regulation of critical cellular processes in cancer. In contrast, MST1R/RON gen is in tumor progression of various types of cancer, and GC mutations have been reported (59).

### Conclusions

The frequency of LOH in specific regions of chromosomes suggests that these loci contain critical regions for tumor suppression or progression. The loss of regions where the sequences of master molecules controlling gene expression (coding and non-coding) are located plays a role in controlling the cell cycle and apoptosis. In contrast, the loss of genes can affect cell adhesion and signaling. These findings support the hypothesis that LOH contributes to genetic instability and GC progression. LOH is an essential mechanism in the oncogenesis of GC that should continue to be investigated. Identifying LOH-genes could provide new opportunities for developing targeted therapeutic strategies and improving the prognosis of patients with GC. Future studies should focus on the functional characterization of these genes and the development of methods to prevent or reverse LOH in precancerous and cancerous cells.

## Acknowledgements

The authors would like to thank Mr. Brian-Alexander Cruz-Ramírez and Ms. Alejandra García-Bejarano, Oncological Diseases Medical Research Unit (UIMEO), UMAE-Oncology Hospital, XXI Century National Medical Center, IMSS for their technical assistance.

## Availability of data and materials

All datasets used and/or analyzed during the current study are available from the corresponding author upon reasonable request. The data have been deposited in the Gene Expression Omnibus Database (GEO; https://www.ncbi.nlm.nih.gov/geo/) under accession no. GSE117093 and BioProjet PRJNA481039.

## Authors’ contributions

VLS participated in data analysis to provide the results, and prepared, wrote, and discussed the manuscript. HAVS performed the molecular experiments and discussed the manuscript. JT, MCP, and PPS were responsible for clinical aspects and recruited the patients. JT, MCP, PPS, and FMS discussed the data and revised the manuscript. MERT contributed to the design of the study, supervised the study, and critically reviewed, revised, and wrote the manuscript. JT and MERT were responsible for acquiring financial support. VLS and MERT confirm the authenticity of all the raw data. All authors read and approved the final manuscript.

## Ethics approval and consent to participate

Institutional Review Board approval was obtained for the study (approval no. 2008-785-001). Clinical data and patient samples were processed following written informed consent.

## Patient consent for publication

Not applicable.

## References

1. Bray F, Laversanne M, Sung H, Ferlay J, Siegel RL, Soerjomataram I and Jemal A: Global cancer statistics 2022: GLOBOCAN estimates of incidence and mortality worldwide for 36 cancers in 185 countries. CA Cancer J Clin 74: 229–263, 2024.

2. Machlowska J, Baj J, Sitarz M, Maciejewski R and Sitarz R: Gastric Cancer: Epidemiology, Risk Factors, Classification, Genomic Characteristics and Treatment Strategies. Int J Mol Sci 21, 2020.

3. Cancer Genome Atlas Research Network: Comprehensive molecular characterization of gastric adenocarcinoma. Nature 513: 202–209, 2014.

4. Lengauer C, Kinzler KW and Vogelstein B: Genetic instabilities in human cancers. Nature 396: 643–649, 1998.

5. Ottini L, Falchetti M, Lupi R, Rizzolo P, Agnese V, Colucci G, Bazan V and Russo A: Patterns of genomic instability in gastric cancer: clinical implications and perspectives. Ann Oncol 17: 97–102, 2006.

6. Liu X and Meltzer SJ: Gastric cancer in the era of precision medicine. Cell Mol Gastroenterol Hepatol 3: 348–358, 2017.

7. Nobili S, Bruno L, Landini I, Napoli C, Bechi P, Tonelli F, Rubio CA, Mini E and Nesi G: Genomic and genetic alterations influence the progression of gastric cancer. World J Gastroenterol 17: 290–299, 2011.

8. Fitzgibbon J, Smith LL, Raghavan M, Smith ML, Debernardi S, Skoulakis S, Lillington D, Lister TA and Young BD: Association between acquired uniparental disomy and homozygous gene mutation in acute myeloid leukemias. Cancer Res 65: 9152–9154, 2005.

9. Ryland GL, Doyle MA, Goode D, Boyle SE, Choong DY, Rowley SM, Li J; Australian Ovarian Cancer Study Group; Bowtell DD, Tothill RW, et al: Loss of heterozygosity: what is it good for? BMC Med Genomics 8: 45, 2015.

10. Tapial S, García JL, Corchete L, Holowatyj AN, Pérez J, Rueda D, Urioste M, González-Sarmiento R and Perea J: Copy neutral loss of heterozygosity (cnLOH) patterns in synchronous colorectal cancer. Eur J Hum Genet 29: 709–713, 2021.

11. Huo X, Xiao X, Zhang S, Du X, Li C, Bai Z and Chen Z: Characterization and clinical evaluation of microsatellite instability and loss of heterozygosity in tumor-related genes in gastric cancer. Oncol Lett 21: 430, 2021.

12. Xiao Y-P, Wu D-Y, Xu L and Xin Y: Loss of heterozygosity and microsatellite instabilities of fragile histidine triad gene in gastric carcinoma. World J Gastroenterol 12: 3766–3769, 2006.

13. Milne AN, Leguit R, Corver WE, Morsink FH, Polak M, de Leng WW, Carvalho R and Offerhaus GJ: Loss of CDC4/FBXW7 in gastric carcinoma. Cell Oncol 32: 347–359, 2010.

14. Zhu X-Y, Yang J-Y, He Y, Liu G-H, Sun Y and Ding Y: Correlation between gastric carcinoma and ZAC gene-associated microsatellite instability and loss of heterozygosity. Oncol Lett 14: 2422–2426, 2017.

15. Chung WC, Jung SH, Lee KM, Paik CN, Kwak JW, Jung JH, Yoo JY, Lee MK and Chung IS: Genetic instability in gastric epithelial neoplasias categorized by the revised vienna classification. Gut Liver 4: 179–185, 2010.

16. Ohshima K, Fujiya K, Nagashima T, Ohnami S, Hatakeyama K, Urakami K, Naruoka A, Watanabe Y, Moromizato S, Shimoda Y, et al: Driver gene alterations and activated signaling pathways toward malignant progression of gastrointestinal stromal tumors. Cancer Sci 110: 3821–3833, 2019.

17. Arakawa N, Sugai T, Habano W, Eizuka M, Sugimoto R, Akasaka R, Toya Y, Yamamoto E, Koeda K, Sasaki A, et al: Genome-wide analysis of DNA copy number alterations in early and advanced gastric cancers. Mol Carcinog 56: 527–537, 2017.

18. Choi WH, Lee S and Cho S: Microsatellite Alterations and Protein Expression of 5 Major Tumor Suppressor Genes in Gastric Adenocarcinomas. Transl Oncol 11: 43–55, 2018.

19. Choi JH, Kim YB, Ahn JM, Kim MJ, Bae WJ, Han SU, Woo HG and Lee D: Identification of genomic aberrations associated with lymph node metastasis in diffuse-type gastric cancer. Exp Mol Med 50: 1–11, 2018.

20. Wang H, Xu M, Cui X, Liu Y, Zhang Y, Sui Y, Wang D, Peng L, Wang D and Yu J: Aberrant expression of the candidate tumor suppressor gene DAL-1 due to hypermethylation in gastric cancer. Sci Rep 6: 21755, 2016.

21. Battista S, Ambrosio MR, Limarzi F, Gallo G and Saragoni L: Molecular alterations in gastric preneoplastic lesions and early gastric cancer. Int J Mol Sci 22, 2021.

22. Wu X, Wang Q, Liu P, Sun L and Wang Y: Potential value of the homologous recombination deficiency signature we developed in the prognosis and drug sensitivity of gastric cancer. Front Genet 13: 1026871, 2022.

23. Hosseini HA, Ahani A, Galehdari H, Froughmand AM, Hosseini M, Masjedizadeh A and Zali MR: Frequent loss of heterozygosity at 8p22 chromosomal region in diffuse type of gastric cancer. World J Gastroenterol 13: 3354–3358, 2007.

24. Larios-Serrato V, Martínez-Ezquerro J-D, Valdez-Salazar H-A, Torres J, Camorlinga-Ponce M, Piña-Sánchez P and Ruiz-Tachiquín M-E: Copy number alterations and epithelial-mesenchymal transition genes in diffuse and intestinal gastric cancers in Mexican patients. Mol Med Rep 25:191, 2022.

25. Wall L, Christiansen T and Orwant J: Programming Perl. 3^rd^ (ed.) O’Reilly Media, Incorporated, California, pp 1–1067, 2000.

26. Bardou P, Mariette J, Escudié F, Djemiel C and Klopp C: jvenn: an interactive Venn diagram viewer. BMC Bioinformatics 15: 293, 2014.

27. Sondka Z, Dhir NB, Carvalho-Silva D, Jupe S, Madhumita, McLaren K, Starkey M, Ward S, Wilding J, Ahmed M, et al: COSMIC: a curated database of somatic variants and clinical data for cancer. Nucleic Acids Res 52: D1210–D1217, 2024.

28. Liang P-I, Wang C-C, Cheng H-J, Wang S-S, Lin Y-C, Lin P and Tung C-W: Curation of cancer hallmark-based genes and pathways for in silico characterization of chemical carcinogenesis. Database 2020:baaa045, 2020.

29. Fabregat A, Jupe S, Matthews L, Sidiropoulos K, Gillespie M, Garapati P, Haw R, Jassal B, Korninger F, May B, et al: The Reactome Pathway Knowledgebase. Nucleic Acids Res 46: D649–D655, 2018.

30. Szklarczyk D, Gable AL, Nastou KC, Lyon D, Kirsch R, Pyysalo S, Doncheva NT, Legeay M, Fang T, Bork P, et al: The STRING database in 2021: customizable protein-protein networks, and functional characterization of user-uploaded gene/measurement sets. Nucleic Acids Res 49: D605–D612, 2021.

31. Shannon P, Markiel A, Ozier O, Baliga NS, Wang JT, Ramage D, Amin N, Schwikowski B and Ideker T: Cytoscape: a software environment for integrated models of biomolecular interaction networks. Genome Res 13: 2498–2504, 2003.

32. Koo SH, Jeong TE, Kang J, Kwon KC, Park JW and Noh SM: Prognostic implications for gastric carcinoma based on loss of heterozygosity genotypes correlation with clinicopathologic variables. Cancer Genet Cytogenet 153: 26–31, 2004.

33. Hwang MS, Mog BJ, Douglass J, et al: Targeting loss of heterozygosity for cancer-specific immunotherapy. Proc Natl Acad Sci U S A 118:e2022410118, 2021.

34. Bellini MF, Cadamuro ACT, Succi M, Proença MA and Silva AE: Alterations of the TP53 gene in gastric and esophageal carcinogenesis. J Biomed Biotechnol 2012: 891961, 2012.

35. Li Y-L, Tian Z, Wu D-Y, Fu B-Y and Xin Y: Loss of heterozygosity on 10q23.3 and mutation of tumor suppressor gene PTEN in gastric cancer and precancerous lesions. World J Gastroenterol 11: 285–288, 2005.

36. Carvalho R, Milne ANA, Polak M, Corver WE, Offerhaus GJA and Weterman MAJ: Exclusion of RUNX3 as a tumour-suppressor gene in early-onset gastric carcinomas. Oncogene 24: 8252–8258, 2005.

37. Poremba C, Yandell DW, Huang Q, Little JB, Mellin W, Schmid KW, Böcker W and Dockhorn-Dworniczak B: Frequency and spectrum of p53 mutations in gastric cancer--a molecular genetic and immunohistochemical study. Virchows Arch 426: 447–455, 1995.

38. Oki E, Tokunaga E, Nakamura T, Ueda N, Futatsugi M, Mashino K, Yamamoto M, Watanabe M, Ikebe M, Kakeji Y, et al: Genetic mutual relationship between PTEN and p53 in gastric cancer. Cancer Lett 227: 33–38, 2005.

39. Neufeld G, Mumblat Y, Smolkin T, Toledano S, Nir-Zvi I, Ziv K and Kessler O: The role of the semaphorins in cancer. Cell Adh Migr 10: 652–674, 2016.

40. Kobayashi T, Chanmee T and Itano N: Hyaluronan: Metabolism and Function. Biomolecules 10:1525, 2020.

41. Vizoso FJ, del Casar JM, Corte MD, García I, Corte MG, Alvarez A and García-Muñiz JL: Significance of cytosolic hyaluronan levels in gastric cancer. Eur J Surg Oncol 30: 318–324, 2004.

42. Liu M, Que Y, Hong Y, Zhang L, Zhang X and Zhang Y: A pan-cancer analysis of IRAK1 expression and their association with immunotherapy response. Front Mol Biosci 9: 904959, 2022.

43. Yin M, Wang X and Lu J: Advances in IKBKE as a potential target for cancer therapy. Cancer Med 9: 247–258, 2020.

44. Gong Y, Zhao W, Jia Q, Dai J, Chen N, Chen Y, Gu D, Huo X and Chen J: IKBKB rs2272736 is associated with gastric cancer survival. Pharmgenomics Pers Med 13: 345– 352, 2020.

45. Wang G-J, Yu T-Y, Li Y-R, Liu Y-J and Deng B-B: Circ_0000190 suppresses gastric cancer progression potentially via inhibiting miR-1252/PAK3 pathway. Cancer Cell Int 20: 351, 2020.

46. Kämmerer U, Gires O, Pfetzer N, Wiegering A, Klement RJ and Otto C: TKTL1 expression in human malign and benign cell lines. BMC Cancer 15: 2, 2015.

47. Ahopelto K, Laitinen A, Hagström J, Böckelman C and Haglund C: Transketolase-like protein 1 and glucose transporter 1 in gastric cancer. Oncology 98: 643–652, 2020.

48. Wang D, Chen Y, Fang H, Zheng L, Li Y, Yang F, Xu Y, Du L, Zhou BS and Li H: Increase of PRPP enhances chemosensitivity of PRPS1 mutant acute lymphoblastic leukemia cells to 5-fluorouracil. J Cell Mol Med 22: 6202–6212, 2018.

49. Feichtinger RG, Neureiter D, Skaria T, Wessler S, Cover TL, Mayr JA, Zimmermann FA, Posselt G, Sperl W and Kofler B: Oxidative phosphorylation system in gastric carcinomas and gastritis. Oxid Med Cell Longev 2017: 1320241, 2017.

50. Puurand M, Peet N, Piirsoo A, Peetsalu M, Soplepmann J, Sirotkina M, Peetsalu A, Hemminki A and Seppet E: Deficiency of the complex I of the mitochondrial respiratory chain but improved adenylate control over succinate-dependent respiration are human gastric cancer-specific phenomena. Mol Cell Biochem 370: 69–78, 2012.

51. Du Y and Grandis JR: Receptor-type protein tyrosine phosphatases in cancer. Chin J Cancer 34: 61–69, 2015.

52. Holsinger LJ, Ward K, Duffield B, Zachwieja J and Jallal B: The transmembrane receptor protein tyrosine phosphatase DEP1 interacts with p120(ctn). Oncogene 21: 7067–7076, 2002.

53. Xu S and Powers MA: Nuclear pore proteins and cancer. Semin Cell Dev Biol 20: 620– 630, 2009.

54. Nam S, Kim JH and Lee DH: RHOA in Gastric Cancer: Functional Roles and Therapeutic Potential. Front Genet 10: 438, 2019.

55. Kaibuchi K, Kuroda S and Amano M: Regulation of the cytoskeleton and cell adhesion by the Rho family GTPases in mammalian cells. Annu Rev Biochem 68: 459–486, 1999.

56. Li M, Lu M, Li J, Gui Q, Xia Y, Lu C and Shu H: Single-cell data revealed CD14-type and FCGR3A-type macrophages and relevant prognostic factors for predicting immunotherapy and prognosis in stomach adenocarcinoma. PeerJ 12: e16776, 2024.

57. Cazes A, Childers BG, Esparza E and Lowy AM: The MST1R/RON tyrosine kinase in cancer: Oncogenic functions and therapeutic strategies. Cancers 14: 2037, 2022.

58. Niloofa R, De Zoysa MI and Seneviratne LS: Autoantibodies in the diagnosis, prognosis, and prediction of colorectal cancer. J Cancer Res Ther 17: 819–833, 2021.

59. Purwar R, Tripathi M, Rajput M, Pal M and Pandey M: Novel mutations in a second primary *gastric* cancer in a patient treated for primary colon cancer. World J Surg Oncol 21: 173, 2023.

